# Efficiency of student counseling among Japanese university students: Comparisons based on sex, therapy status, and the number of sessions attended

**DOI:** 10.1101/2025.11.16.25340355

**Authors:** Ryo Horita, Cathelencia Francisque, Nicole Nagib, Taku Fukao, Nanako Imamura, Yumiko Kuriki, Ryoko Okamoto, Mayumi Yamamoto

## Abstract

University students worldwide face increasing mental health challenges. Yet, research on the effectiveness of campus-based counseling in Japan is limited. This study evaluated changes in psychological symptoms among Japanese university students receiving counseling, using the Counseling Center Assessment of Psychological Symptoms—Japanese version (CCAPS-Japanese). Data were collected from 99 students at a national Japanese university who voluntarily sought counseling between November 2021 and July 2024. They completed the CCAPS-Japanese pre- and post-counseling. Outcomes were analyzed using paired *t*-tests and effect sizes across sex, therapy status (terminated or interrupted), and number of sessions attended (short-term [2 sessions], mid-term [3–10 sessions], or long-term [≥ 11 sessions]). Students showed significant improvements in depression, generalized anxiety, thought disturbances, and suicidal ideation. Effect sizes ranged from moderate to large. Improvements were noted regardless of sex or therapy status, although sex-specific patterns emerged: male students showed improvements in social anxiety and depression, whereas female students showed improvements in hostility. Both the terminated and interrupted groups exhibited significant symptom reductions; however, only the terminated group improved in academic distress. Furthermore, mid-term counseling yielded the most consistent improvements, whereas short-term counseling yielded no significant changes. Counseling effectively reduces key psychological symptoms among Japanese university students, particularly depression and anxiety. Mid-term interventions appear most beneficial for achieving therapeutic impact. These findings empirically support university counseling services in Japan and highlight the need for continued investment in and outcome monitoring of student mental health programs.

## Introduction

University often represents a pivotal life stage, during which young adults navigate independence and new responsibilities for the first time [1]. This transition can be a stressful time, as it entails substantial changes. Consequently, psychological distress can be a part of the university experience. Notably, during the 2020–2021 academic year, over 60% of US students met the criteria for at least one mental health condition [2]. University students face a range of psychological challenges, including depression, anxiety, suicidal ideation, eating disorders, and substance misuse [3]. Such difficulties are often exacerbated by academic pressure, financial concerns, and the transitional stressors associated with emerging adulthood. Moreover, emotional stress and mental health concerns have been cited as a leading cause of attrition in higher education [4]. Consequently, student mental health crises in higher education are increasingly recognized as systemic issues requiring institutional and societal responses.

To address these challenges, many universities have established counseling centers designed to support students’ mental health and academic success. Evidence indicates that these centers are effective in managing mild mental illnesses [5]. Most students who seek counseling are self-motivated to get better, presenting with acute concerns that, while urgent, typically do not pose a risk to their safety. Collins et al. [6] evaluated university counseling services using standardized outcome measures and found consistent improvements in psychological functioning among students who received counseling. They emphasized the importance of using routine outcome measures to demonstrate clinical effectiveness and support evidence-based practices in higher education settings. Furthermore, early counseling interventions improve academic performance and reduce interpersonal conflicts among students experiencing psychological symptoms [7,8].

However, research remains scant on the effectiveness of university counseling services in Japan. While several studies have examined the prevalence and trends of mental health challenges among general university students [9,10], few have investigated the therapeutic outcomes for those who utilized counseling services. Although some studies have explored student needs and general mental health trends, comprehensive evaluations of counseling outcomes remain scarce [11,12]. Egami et al. [13] noted that some counselors in Japanese universities perceive evaluating the effectiveness of counseling as difficult and hold negative attitudes toward such evaluations. Cultural factors, including stigma surrounding mental health and a collectivist orientation, may also contribute to counseling underutilization and the underreporting of its effectiveness [14].

These challenges are compounded by the broader context of financial strain within Japanese higher education. As Japan faces demographic decline, many universities are experiencing budget cuts and staff reductions [15]. These constraints threaten the sustainability of student support services, including counseling. Demonstrating the effectiveness of counseling programs is, therefore, critical not only academically but also for institutional accountability and resource allocation.

This study examined whether students who received counseling at a university counseling center in Japan demonstrated improvements in mental health from pre- to post-treatment. It also clarified whether outcomes vary according to sex, therapy status (terminated vs. interrupted), and the number of sessions attended. To the best of our knowledge, no existing studies have systematically evaluated the effectiveness of counseling provided at university counseling centers in Japan. Furthermore, in university counseling centers in the United States, insurance system constraints often limit the number of sessions that can be availed. Consequently, many studies have investigated whether short-term counseling can still be effective. However, Japanese universities—including the institution targeted in this study—generally do not impose restrictions on the number of counseling sessions one can avail. This provides a unique opportunity to assess the effects of long-term interventions, thereby strengthening the contribution of this study to both domestic and international scholarship.

## Methods

### Design, participants, and procedures

We targeted students who used the counseling center at X University, a moderately sized national general university in Japan, between November 26, 2021 and July 31, 2024. The inclusion criteria were: (1) first-time counseling-service users whose therapy was terminated or interrupted between November 2021 and July 2024; (2) completed the Counseling Center Assessment of Psychological Symptoms—Japanese questionnaire; and (3) identified oneself as male or female. Students were excluded if they had previously used the counseling center services, declined to complete the questionnaire at each visit, or identified their sex as “other.”

An online survey [16] was administered at the counseling center’s reception desk immediately before each counseling appointment. All students who visited the counseling center and met the eligibility criteria were invited to participate. Participation was voluntary, and informed consent was obtained before completing the assessment. Students were assured that their decision to participate would have no impact on their academic standing, grades, or access to counseling services.

The counseling sessions were scheduled for 45 minutes, although some lasted slightly longer. The primary intervention consisted of psychotherapy, delivered based on the individual professional style of the four available student counselors. All counselors were licensed as Clinical Psychologists and Certified Public Psychologists in Japan. No specific therapy was recommended for this study.

The therapy status was assessed by each respective counselor in charge. Terminated therapy denoted that the student and their counselor had mutually agreed to end treatment, whereas interrupted therapy denoted that the student had prematurely discontinued attendance without such an agreement. Each student attended a minimum of two sessions, with no upper limit on the number of sessions attended.

Table 1 summarizes participants’ background characteristics. A total of 99 students participated, with 64 male and 35 female students and a mean age of 21.04 years (standard deviation = 2.02). Regarding academic level, 79 participants (79.8%) were undergraduates, and 20 (20.2%) were enrolled in a master’s program. Therapy status was nearly evenly distributed, with slightly more participants in the terminated group (n = 50, 50.5%) than in the interrupted group (n = 49, 49.5%). Regarding the number of sessions attended, the short-term group (2 sessions) was the smallest (n = 22, 22.2%), the mid-term group (3–10 sessions) was the largest (n = 46, 46.5%), and the long-term group (≥ 11 sessions) was the second largest (n = 31, 31.3%). Demographic data (sex, age, and academic level) were collected through an online survey [16]. All data were anonymized and de-identified prior to analysis.

**Table 1.**
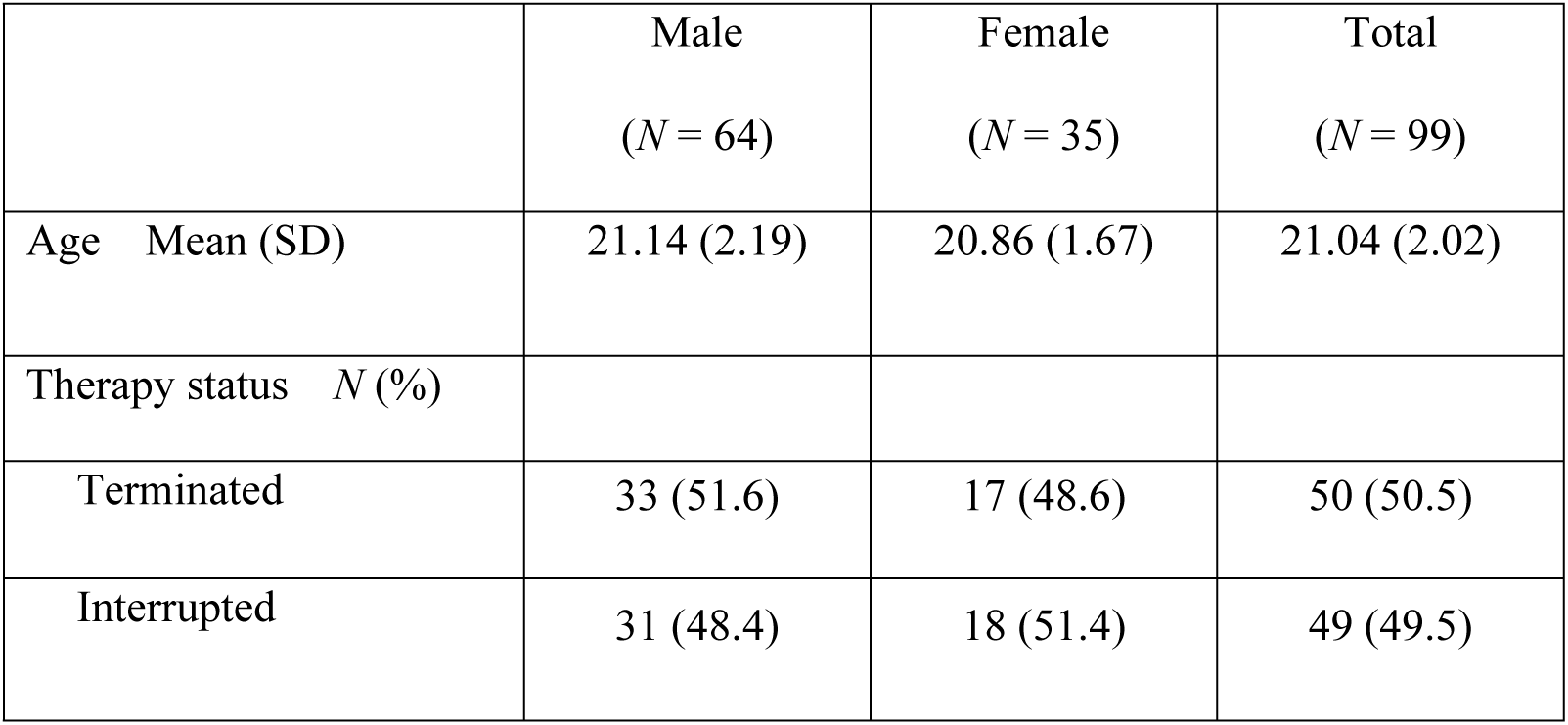
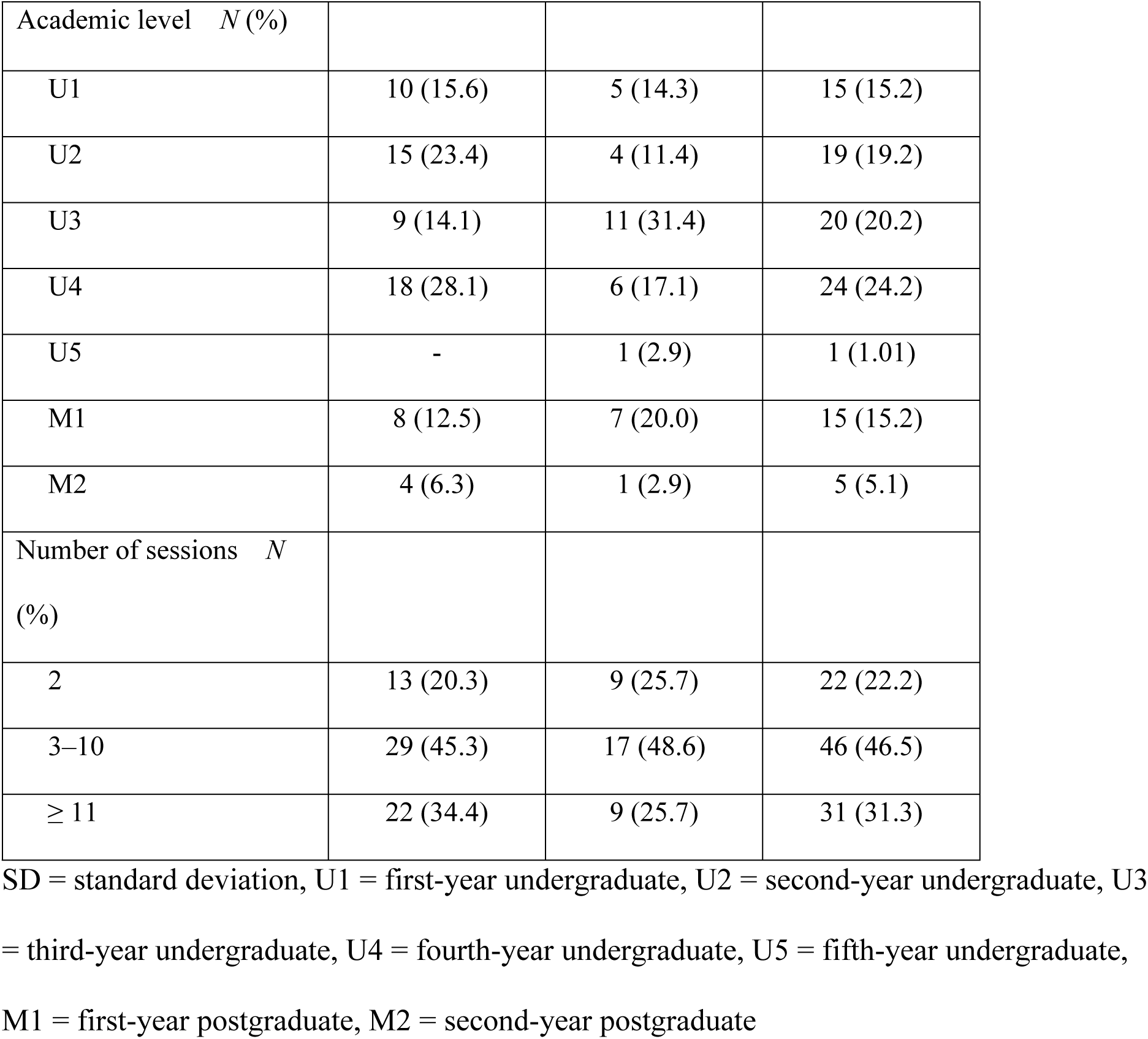
Participants’ background characteristics based on sex.

### Measure

The Counseling Center Assessment of Psychological Symptoms—Japanese version (CCAPS-Japanese [17,18]) was used to assess psychological distress. The CCAPS was originally developed in 2001 by the University of Michigan’s Counseling and Psychological Services to assess psychological symptoms among university students [19]. The CCAPS-Japanese was adapted and validated for use among Japanese university students, demonstrating a robust factor structure, good internal consistency, acceptable convergent validity, and stable test–retest reliability [17,18]. The instrument comprises 55 items across eight subscales: depression (11 items), generalized anxiety (9 items), social anxiety (6 items), academic distress (3 items), eating concerns (8 items), hostility (7 items), family distress (6 items), and alcohol use (5 items). Additionally, it includes four critical items to detect acute risks: thought disturbance (“I lose touch with reality”), suicidal ideation (“I have thoughts of ending my life”), violent behavior (“I am afraid I may lose control and act violently”), and homicidal behavior (“I have thoughts of hurting others”). Participants rate the degree to which each item reflects their experience over the past two weeks on a 5-point Likert scale ranging from 0 (*not at all like me*) to 4 (*extremely like me*), with higher scores indicating greater levels of psychological distress.

### Data analysis

We employed a cohort design to compare pre- and post-intervention scores on the subscales and critical items of the CCAPS-Japanese. The pre-intervention scores were CCAPS-Japanese scores measured immediately before the initial counseling session, while post-intervention scores were CCAPS-Japanese scores measured at the final session for the termination group and at the last attended session for the interruption group. A decrease in post-intervention scores relative to pre-intervention scores indicated greater counseling effectiveness. Scores were analyzed using paired *t*-tests to assess the effect of therapy on psychological symptoms. Analyses were conducted for the entire sample as well as stratified by sex (male or female), therapy status (terminated or interrupted), and number of sessions attended (2 sessions, 3–10 sessions, or ≥ 11 sessions). Statistical significance was defined as *p* < 0.05. Effect sizes were calculated using Cohen’s *d*, interpreted as follows: 0.0 to < 0.2, “little to no effect”; 0.2 to < 0.5, “small effect”; 0.5 to < 0.8, “moderate effect”; and ≥ 0.8, “large effect.” All statistical analyses were performed using Statistical Package for Social Sciences (SPSS) Software version 30 (IBM, Armonk, NY, USA).

### Ethical considerations

The study protocol was approved by the Ethical Review Committee of the Graduate School of Medicine, Gifu University, Japan (Approval Number: 2021-B114). All students received a written informed consent form stating that their data would remain confidential and anonymous. They were informed that participation was voluntary, with no compensation or research credit provided, and that their responses would not affect their academic evaluations. Participants were also advised that they could withdraw from the study at any time without penalty.

## Results

### Results for all participants

Table 2 summarizes the results of paired *t*-tests and effect sizes for CCAPS-Japanese subscales and critical items. Post-intervention scores were significantly lower than pre-intervention scores for the depression (*t*(98) = −3.33, *p* < 0.001, *d* = 0.74) and generalized anxiety (*t*(98) = −3.08, *p* < 0.001, *d* = 0.73) subscales, indicating a moderate effect size. Additionally, post-intervention scores for thought disturbance (*t*(98) = −2.18, *p* = 0.003, *d* = 1.31) and suicidal ideation (*t*(98) = −3.83, *p* < 0.001, *d* = 1.17) were significantly lower than pre-intervention scores, indicating a large effect size. While other CCAPS-Japanese subscales and critical items did not show statistically significant results (all *p* > 0.05), they exhibited substantial effect sizes (Cohen’s *d* = 0.57–1.07).

**Table 2.** Pre- and post-intervention scores and effect sizes for CCAPS-Japanese subscales and critical items.

| Subscale/Critical item | Pre-intervention | Post-intervention | $t$ | Cohen's<br>$d$ | $p$ |
| --- | --- | --- | --- | --- | --- |

|  | score | score |  |  |  |
| --- | --- | --- | --- | --- | --- |
|  | Mean (SD) | Mean (SD) |  |  |  |
| **Subscales** |  |  |  |  |  |
| Depression | 1.99 (0.80) | 1.75 (0.87) | −3.33 | 0.74 | <0.001* |
| Generalized anxiety | 1.93 (0.80) | 1.72 (0.84) | −3.08 | 0.73 | 0.001* |
| Social anxiety | 2.41 (0.87) | 2.35 (0.94) | −1.14 | 0.57 | 0.129 |
| Academic distress | 2.25 (1.10) | 2.14 (1.02) | −1.26 | 0.92 | 0.105 |
| Eating concerns | 1.13 (0.79) | 1.13 (0.83) | 0.15 | 0.61 | 0.439 |
| Hostility | 1.08 (0.83) | 0.98 (0.93) | −1.43 | 0.73 | 0.078 |
| Family distress | 1.20 (0.91) | 1.25 (0.87) | 0.59 | 0.65 | 0.280 |
| Alcohol use | 0.43 (0.70) | 0.51 (0.76) | 1.23 | 0.64 | 0.104 |
| **Critical items** |  |  |  |  |  |
| Thought disturbance | 1.88 (1.37) | 1.48 (1.24) | −2.18 | 1.31 | 0.003* |
| Suicidal ideation | 1.99 (1.27) | 1.57 (1.36) | −3.83 | 1.17 | <0.001* |
| Violent behavior | 0.71 (1.12) | 0.69 (1.07) | −0.19 | 1.07 | 0.426 |
| Homicidal behavior | 0.70 (1.09) | 0.62 (0.98) | −0.80 | 1.00 | 0.213 |

Tested using paired *t*-test. CCAPS-Japanese: The Counseling Center Assessment of Psychological
Symptoms—Japanese, SD: standard deviation. The mean difference was obtained by subtracting the
post-intervention score from the pre-intervention score.
\**p* < 0.05.

### Results based on sex

Table 3 summarizes the results of paired *t*-tests and effect sizes for CCAPS-Japanese subscales and critical items stratified by sex. The post-intervention score for the generalized anxiety subscale was significantly lower in both men (*t*(63) = −2.06, *p* = 0.022, *d* = 0.72) and women (*t*(34) = −2.03, *p* = 0.025, *d* = 0.73) compared with the pre-intervention score, indicating moderate effect sizes. Depression (*t*(63) = −2.79, *p* = 0.004, *d* = 0.77) and social anxiety (*t*(63) = −2.37, *p* = 0.010, *d* = 0.55) decreased significantly only in men, whereas hostility decreased significantly only in women (*t*(34) = −1.78, *p* = 0.042, *d* = 0.79), all reflecting moderate effect sizes. Furthermore, thought disturbance and suicidal ideation scores decreased significantly in men (*t*(63) = −2.23, *p* = 0.015, *d* = 1.29 and *t*(63) = −3.07, *p* = 0.002, *d* = 1.18, respectively) as well as women (*t*(34) = −2.02, *p* = 0.025, *d* = 1.34 and *t*(34) = −1.89, *p* = 0.034, *d* = 1.17, respectively), indicating large effect sizes.

**Table 3.**
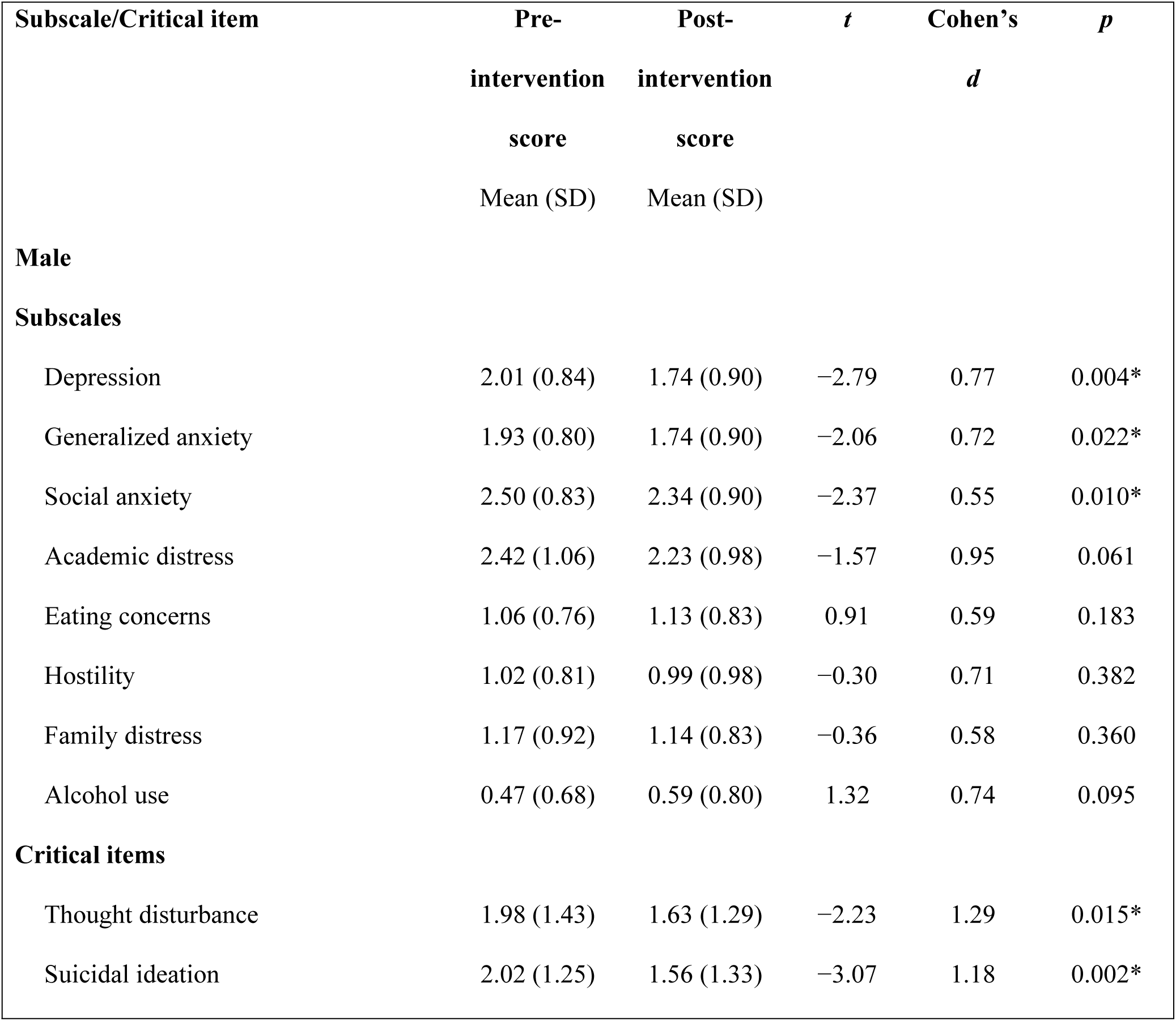
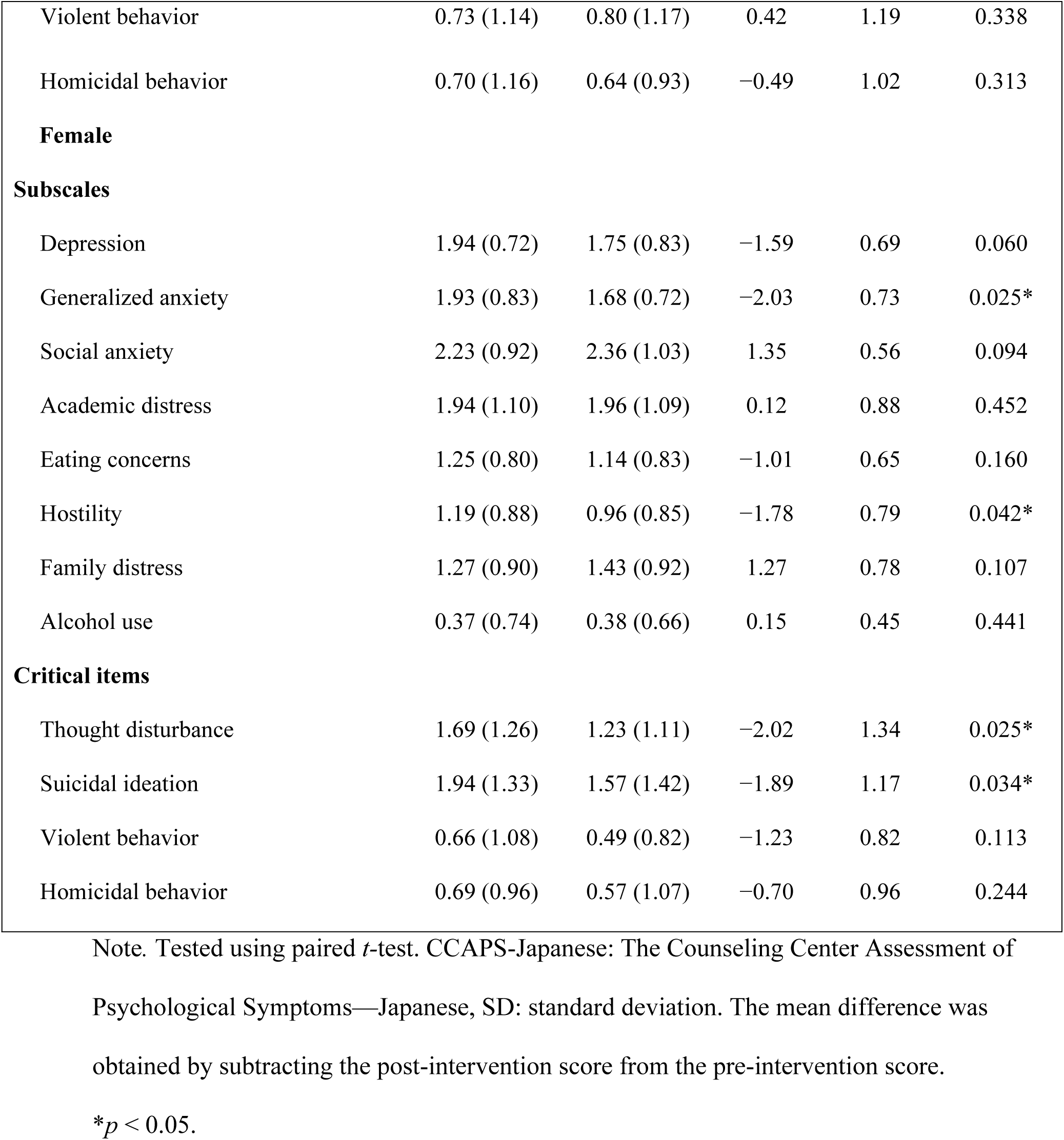
Pre- and post-intervention scores and effect sizes for CCAPS-Japanese subscales and critical items stratified based on sex.

| Subscale/Critical item | Pre-intervention<br>score<br>Mean (SD) | Post-intervention<br>score<br>Mean (SD) | *t* | Cohen's<br>*d* | *p* |
| --- | --- | --- | --- | --- | --- |
| **Male** |  |  |  |  |  |
| **Subscales** |  |  |  |  |  |
| Depression | 2.01 (0.84) | 1.74 (0.90) | -2.79 | 0.77 | 0.004* |
| Generalized anxiety | 1.93 (0.80) | 1.74 (0.90) | -2.06 | 0.72 | 0.022* |
| Social anxiety | 2.50 (0.83) | 2.34 (0.90) | -2.37 | 0.55 | 0.010* |
| Academic distress | 2.42 (1.06) | 2.23 (0.98) | -1.57 | 0.95 | 0.061 |
| Eating concerns | 1.06 (0.76) | 1.13 (0.83) | 0.91 | 0.59 | 0.183 |
| Hostility | 1.02 (0.81) | 0.99 (0.98) | -0.30 | 0.71 | 0.382 |
| Family distress | 1.17 (0.92) | 1.14 (0.83) | -0.36 | 0.58 | 0.360 |
| Alcohol use | 0.47 (0.68) | 0.59 (0.80) | 1.32 | 0.74 | 0.095 |
| **Critical items** |  |  |  |  |  |
| Thought disturbance | 1.98 (1.43) | 1.63 (1.29) | -2.23 | 1.29 | 0.015* |
| Suicidal ideation | 2.02 (1.25) | 1.56 (1.33) | -3.07 | 1.18 | 0.002* |
| Violent behavior | 0.73 (1.14) | 0.80 (1.17) | 0.42 | 1.19 | 0.338 |
| Homicidal behavior | 0.70 (1.16) | 0.64 (0.93) | −0.49 | 1.02 | 0.313 |
| **Female** |  |  |  |  |  |
| **Subscales** |  |  |  |  |  |
| Depression | 1.94 (0.72) | 1.75 (0.83) | −1.59 | 0.69 | 0.060 |
| Generalized anxiety | 1.93 (0.83) | 1.68 (0.72) | −2.03 | 0.73 | 0.025* |
| Social anxiety | 2.23 (0.92) | 2.36 (1.03) | 1.35 | 0.56 | 0.094 |
| Academic distress | 1.94 (1.10) | 1.96 (1.09) | 0.12 | 0.88 | 0.452 |
| Eating concerns | 1.25 (0.80) | 1.14 (0.83) | −1.01 | 0.65 | 0.160 |
| Hostility | 1.19 (0.88) | 0.96 (0.85) | −1.78 | 0.79 | 0.042* |
| Family distress | 1.27 (0.90) | 1.43 (0.92) | 1.27 | 0.78 | 0.107 |
| Alcohol use | 0.37 (0.74) | 0.38 (0.66) | 0.15 | 0.45 | 0.441 |
| **Critical items** |  |  |  |  |  |
| Thought disturbance | 1.69 (1.26) | 1.23 (1.11) | −2.02 | 1.34 | 0.025* |
| Suicidal ideation | 1.94 (1.33) | 1.57 (1.42) | −1.89 | 1.17 | 0.034* |
| Violent behavior | 0.66 (1.08) | 0.49 (0.82) | −1.23 | 0.82 | 0.113 |
| Homicidal behavior | 0.69 (0.96) | 0.57 (1.07) | −0.70 | 0.96 | 0.244 |

Note. Tested using paired *t*-test. CCAPS-Japanese: The Counseling Center Assessment of Psychological Symptoms—Japanese, SD: standard deviation. The mean difference was obtained by subtracting the post-intervention score from the pre-intervention score.
\* $p < 0.05$ .

### Results based on therapy status

Table 4 presents the results of paired *t*-tests and effect sizes for CCAPS-Japanese subscales and critical items stratified by therapy status. Post-intervention depression and generalized anxiety scores decreased significantly in the “terminated” (*t*(49) = −2.13, *p* = 0.019, *d* = 0.75 and *t*(49) = −1.84, *p* = 0.036, *d* = 0.68, respectively) as well as the “interrupted” group (*t*(48) = −2.57, *p* = 0.007, *d* = 0.73 and *t*(48) = −2.48, *p* = 0.008, *d* = 0.77, respectively), indicating moderate effect sizes. In the “terminated” group, academic distress scores decreased significantly (*t*(49) = −2.28, *p* = 0.014, *d* = 0.73), while alcohol use scores increased significantly (*t*(49) = 2.28, *p* = 0.013, *d* = 0.58), both reflecting moderate effects. Furthermore, suicidal ideation scores decreased significantly in both the “terminated” (*t*(49) = −3.07, *p* = 0.002, *d* = 1.01) and “interrupted” groups (*t*(48) = −2.16, *p* = 0.018, *d* = 1.32), indicating large effect sizes. In the “interrupted” group, the post-intervention score for thought disturbance was significantly lower than the pre-intervention score (*t*(48) = −2.73, *p* = 0.004, *d* = 1.31), indicating a large effect size.

**Table 4.** Pre- and post-intervention scores and effect sizes for CCAPS-Japanese subscales and critical items stratified based on therapy status.

| Subscale/Critical item | Pre-intervention<br>score<br>Mean (SD) | Post-intervention<br>score<br>Mean (SD) | $t$ | Cohen's<br>$d$ | $p$ |
| --- | --- | --- | --- | --- | --- |
| **Terminated** |  |  |  |  |  |
| **Subscales** |  |  |  |  |  |
| Depression | 1.88 (0.84) | 1.67 (0.89) | -2.13 | 0.75 | 0.019* |
| Generalized anxiety | 1.82 (0.85) | 1.67 (0.83) | -1.84 | 0.68 | 0.036* |
| Social anxiety | 2.30 (0.94) | 2.21 (0.90) | -1.25 | 0.56 | 0.109 |
| Academic distress | 2.04 (1.10) | 1.81 (1.04) | -2.28 | 0.73 | 0.014* |
| Eating concerns | 1.07 (0.73) | 1.13 (0.82) | 0.88 | 0.59 | 0.192 |
| Hostility | 1.02 (0.84) | 0.91 (0.94) | -1.24 | 0.64 | 0.110 |
| Family distress | 1.22 (0.96) | 1.29 (0.85) | 0.66 | 0.60 | 0.256 |
| Alcohol use | 0.33 (0.67) | 0.52 (0.74) | 2.28 | 0.58 | 0.013* |
| **Critical items** |  |  |  |  |  |
| Thought disturbance | 1.76 (1.51) | 1.48 (1.34) | −1.53 | 1.29 | 0.066 |
| Suicidal ideation | 1.92 (1.31) | 1.48 (1.28) | −3.07 | 1.01 | 0.002* |
| Violent behavior | 0.72 (1.03) | 0.70 (1.11) | −0.14 | 1.02 | 0.445 |
| Homicidal behavior | 0.72 (1.09) | 0.62 (0.92) | −0.82 | 0.86 | 0.208 |
| **Interrupted** |  |  |  |  |  |
| **Subscales** |  |  |  |  |  |
| Depression | 2.09 (0.75) | 1.83 (0.85) | −2.57 | 0.73 | 0.007* |
| Generalized anxiety | 2.04 (0.74) | 1.77 (0.86) | −2.48 | 0.77 | 0.008* |
| Social anxiety | 2.52 (0.78) | 2.49 (0.97) | −0.36 | 0.58 | 0.359 |
| Academic distress | 2.47 (1.05) | 2.48 (0.89) | 0.04 | 1.08 | 0.483 |
| Eating concerns | 1.19 (0.84) | 1.14 (0.84) | −0.62 | 0.64 | 0.270 |
| Hostility | 1.15 (0.83) | 1.05 (0.93) | −0.84 | 0.83 | 0.204 |
| Family distress | 1.18 (0.85) | 1.20 (0.89) | 0.20 | 0.71 | 0.422 |
| Alcohol use | 0.54 (0.72) | 0.51 (0.78) | −0.29 | 0.69 | 0.387 |
| **Critical items** |  |  |  |  |  |
| Thought disturbance | 2.00 (1.23) | 1.49 (1.14) | −2.73 | 1.31 | 0.004* |
| Suicidal ideation | 2.06 (1.25) | 1.65 (1.44) | −2.16 | 1.32 | 0.018* |
| Violent behavior | 0.69 (1.21) | 0.67 (1.03) | −0.13 | 1.15 | 0.451 |
| Homicidal behavior | 0.67 (1.11) | 0.61 (1.04) | −0.38 | 1.13 | 0.353 |

Note. Tested using paired *t*-test. CCAPS-Japanese: The Counseling Center Assessment of
Psychological Symptoms—Japanese, SD: standard deviation. The mean difference was obtained by
subtracting the post-intervention score from the pre-intervention score.
\* $p < 0.05$ .

### Results based on the number of sessions attended

Table 5 shows the results of paired *t*-tests and effect sizes for CCAPS-Japanese subscales and critical items stratified by the number of sessions attended. In the short-term group, no significant differences were observed in the pre- and post-intervention scores for CCAPS-Japanese subscales and critical items. In the mid-term group, depression (*t*(45) = −2.38, *p* = 0.011, *d* = 0.76) and generalized anxiety (*t*(45) = −2.12, *p* = 0.020, *d* = 0.78) decreased significantly, reflecting moderate effect sizes. Similarly, the long-term group showed reductions in depression (*t*(30) = −1.88, *p* = 0.035, *d* = 0.84) and generalized anxiety (*t*(30) = −1.78, *p* = 0.042, *d* = 0.82). However, eating concerns increased significantly in the long-term group (*t*(30) = 2.37, *p* = 0.012, *d* = 0.62), indicating a moderate effect size. Regarding critical items, suicidal ideation decreased significantly in the mid-term (*t*(45) = −2.38, *p* = 0.011, *d* = 1.24) as well as long-term (*t*(30) = −3.23, *p* = 0.001, *d* = 1.11) groups, indicating large effect sizes. Thought disturbance decreased significantly in the mid-term group (*t*(45) = −2.39, *p* = 0.011, *d* = 1.30), also showing a large effect size.

**Table 5.** Pre- and post-intervention scores and effect sizes for CCAPS-Japanese subscales and critical items stratified based on the number of sessions attended.

| Subscale/Critical item | Pre-intervention<br>score<br>Mean (SD) | Post-intervention<br>score<br>Mean (SD) | *t* | Cohen's<br>*d* | *p* |
| --- | --- | --- | --- | --- | --- |
| **Short-term (2 sessions)** |  |  |  |  |  |
| **Subscales** |  |  |  |  |  |
| Depression | 2.10 (0.96) | 1.95 (0.90) | -1.35 | 0.53 | 0.096 |
| Generalized anxiety | 2.06 (0.87) | 1.92 (0.87) | -1.46 | 0.42 | 0.080 |
| Social anxiety | 2.42 (1.04) | 2.41 (1.00) | -0.08 | 0.38 | 0.467 |
| Academic distress | 2.49 (0.99) | 2.32 (1.01) | −1.18 | 0.67 | 0.126 |
| Eating concerns | 1.51 (0.83) | 1.34 (0.98) | −1.56 | 0.53 | 0.067 |
| Hostility | 1.18 (0.76) | 1.15 (0.84) | −0.24 | 0.49 | 0.407 |
| Family distress | 1.00 (0.92) | 1.02 (0.91) | 0.23 | 0.49 | 0.411 |
| Alcohol use | 0.38 (0.60) | 0.41 (0.61) | 0.24 | 0.54 | 0.407 |
| **Critical items** |  |  |  |  |  |
| Thought disturbance | 2.05 (1.29) | 1.68 (1.13) | −1.56 | 1.09 | 0.067 |
| Suicidal ideation | 2.05 (1.50) | 1.95 (1.33) | −0.40 | 1.07 | 0.346 |
| Violent behavior | 0.73 (1.08) | 0.59 (0.96) | −0.57 | 1.13 | 0.288 |
| Homicidal behavior | 0.59 (0.85) | 0.59 (1.05) | 0.00 | 0.82 | 0.500 |
| **Mid-term (3–10 sessions)** |  |  |  |  |  |
| **Subscales** |  |  |  |  |  |
| Depression | 1.86 (0.61) | 1.60 (0.76) | −2.38 | 0.76 | 0.011* |
| Generalized anxiety | 1.75 (0.73) | 1.54 (0.76) | −2.12 | 0.78 | 0.020* |
| Social anxiety | 2.30 (0.90) | 2.26 (0.98) | −0.65 | 0.60 | 0.259 |
| Academic distress | 2.12 (1.08) | 2.04 (1.13) | −0.58 | 0.92 | 0.284 |
| Eating concerns | 1.01 (0.75) | 0.93 (0.73) | −0.81 | 0.60 | 0.210 |
| Hostility | 0.93 (0.76) | 0.81 (0.83) | −1.19 | 0.69 | 0.119 |
| Family distress | 1.28 (0.90) | 1.25 (0.90) | −0.34 | 0.71 | 0.368 |
| Alcohol use | 0.40 (0.66) | 0.43 (0.70) | 0.27 | 0.53 | 0.393 |
| **Critical items** |  |  |  |  |  |
| Thought disturbance | 1.57 (1.23) | 1.11 (1.04) | −2.39 | 1.30 | 0.011* |
| Suicidal ideation | 1.83 (1.12) | 1.39 (1.33) | −2.38 | 1.24 | 0.011* |
| Violent behavior | 0.52 (0.98) | 0.61 (0.95) | 0.66 | 0.89 | 0.255 |
| Homicidal behavior | 0.65 (1.02) | 0.54 (0.94) | −0.73 | 1.02 | 0.236 |
| **Long-term (≥ 11 sessions)** |  |  |  |  |  |
| **Subscales** |  |  |  |  |  |
| Depression | 2.09 (0.91) | 1.81 (0.98) | −1.88 | 0.84 | 0.035* |
| Generalized anxiety | 2.10 (0.82) | 1.85 (0.91) | −1.78 | 0.82 | 0.042* |
| Social anxiety | 2.55 (0.68) | 2.43 (0.86) | −1.03 | 0.64 | 0.156 |
| Academic distress | 2.29 (1.19) | 2.15 (0.86) | −0.71 | 1.09 | 0.240 |
| Eating concerns | 1.03 (0.75) | 1.29 (0.80) | 2.37 | 0.62 | 0.012* |
| Hostility | 1.24 (0.96) | 1.11 (1.10) | −0.83 | 0.93 | 0.207 |
| Family distress | 1.24 (0.91) | 1.40 (0.78) | 1.35 | 0.67 | 0.094 |
| Alcohol use | 0.50 (0.83) | 0.72 (0.91) | 1.40 | 0.85 | 0.086 |
| **Critical items** |  |  |  |  |  |
| Thought disturbance | 2.23 (1.52) | 1.90 (1.45) | −1.22 | 1.47 | 0.116 |
| Suicidal ideation | 2.19 (1.33) | 1.55 (1.41) | −3.23 | 1.11 | 0.001* |
| Violent behavior | 0.97 (1.30) | 0.87 (1.28) | −0.41 | 1.30 | 0.341 |
| Homicidal behavior | 0.84 (1.34) | 0.74 (1.00) | −0.49 | 1.11 | 0.315 |

Note. Tested using paired *t*-test. CCAPS-Japanese: The Counseling Center Assessment of
Psychological Symptoms—Japanese, SD: standard deviation. The mean difference was
obtained by subtracting the post-intervention score from the pre-intervention score.
\* $p < 0.05$ .

## Discussion

This study assessed the effectiveness of university counseling using the CCAPS-Japanese as the psychological questionnaire. Different factors were examined, including sex, therapy status, and the number of sessions attended. Specifically, significant improvements were observed in depression, generalized anxiety, thought disturbance, and suicidal ideation post-counseling. The Center for Collegiate Mental Health [20]—a practice-research network focused on collecting and analyzing mental health data from university and college counseling centers—emphasizes that suicide prevention and depression- and anxiety-related distress reduction are critical roles of counseling centers. Therefore, this study holds particular significance by demonstrating that counseling effectively addresses these critical issues. The findings indicate that the degree of improvement from pre- to post-counseling intervention differs based on sex and between the “terminated” and “interrupted” groups. Additionally, counseling limited to only two sessions may be insufficient to produce meaningful change, whereas three to ten sessions appear to offer the most substantial benefits. These results suggest that ensuring clients receive a sufficient number of sessions—particularly within the range of three to ten—may be crucial for maximizing the effectiveness of counseling interventions.

Notably, the average pre-counseling scores for depression and generalized anxiety subscales among the study participants exceeded the established clinical cutoff points [21], indicating that university students utilizing the counseling center experienced considerable psychological stress. Thus, depression and generalized anxiety appear to be the primary targets for intervention in university counseling settings [20]. The findings of this study consistently demonstrated reductions in depressive and generalized anxiety symptoms, thereby providing empirical support for the efficacy of counseling. In this study, factors such as academic distress, eating concerns, family distress, and alcohol use did not show statistically significant changes. While depression and anxiety are relatively common concerns among students who seek counseling, issues related to academic performance, eating behavior, family relationships, and alcohol dependence may not be shared by all clients. Because this study did not account for the primary complaints of the students who utilized counseling services, different outcomes may emerge if students presenting these specific concerns were identified and analyzed separately. For example, eating concerns did not significantly improve overall, except in the long-term subgroup, which unexpectedly showed an increase following therapy. Although significant, this increase may have limited clinical relevance, as lesser number of sessions showed a nonsignificant decrease in scores. Most students with eating concerns typically tend to avoid counseling, partly due to the secretive nature of these issues [22]. Although studies have shown that psychotherapy is effective for addressing the symptoms of eating disorders [23], treatment approaches must be tailored to the specific type of eating disorder. Scores for alcohol use increased overall; however, the average was between 0 (*not at all like me*) and 1 (*a little like me*), suggesting alcohol-related problems were relatively uncommon among the present sample.

Sex-based differences in therapeutic outcomes were also evident. Lambert [24] analyzed data from more than 17,000 university students who received treatment at university counseling centers and found that female students generally experienced greater therapeutic benefits than their male counterparts. In this study, both male and female students exhibited significant reductions in generalized anxiety, thought disturbance, and suicidal ideation.

Significant improvements in depression and social anxiety were observed only among men, whereas only women exhibited significant reductions in hostility. These findings both support and extend Lambert’s seminal work by replicating overall mental-health change patterns with a Japanese university sample and by further delineating sex-specific trajectories of symptom improvement. Previous research involving Japanese university students has demonstrated that differences in gender role attitudes and dependency needs influence students’ reluctance to seek counseling [25]. Specifically, male students tend to show greater resistance to depending on others and are more likely to view counseling negatively, whereas female students exhibit higher dependency needs and tend to be more receptive to counseling. While these findings are based on the general student population, this study focused specifically on students who availed counseling services. The fact that male students in this study chose to seek counseling despite potential stigma may reflect stronger expectations for the intervention, which could, in turn, have contributed to the greater therapeutic gains observed in this group.

Stratifying results based on therapy status revealed that the “interrupted” group surprisingly still demonstrated significant post-counseling improvements. Students who discontinued their therapy sessions showed changed levels of depression, generalized anxiety, thought disturbance, and suicidal ideation. This finding may indicate that some students benefited sufficiently from the sessions they attended and felt no need to continue. Alternatively, it may represent students who had mild enough disturbances that did not require more sessions. Conversely, in the “terminated” group, the average post-intervention scores for depression and generalized anxiety subscales fell below the clinical cutoff points [21]. This suggests that students who successfully completed their counseling sessions experienced a reduction in depressive and anxiety symptoms to levels unlikely to interfere with their daily functioning. Within this group, depression and generalized anxiety continued to show the greatest improvement, and academic distress also decreased significantly. Seeing that an effective way exists for managing student distress demonstrates the importance of addressing this issue. Notably, academic distress reduced significantly in the “terminated” group but not in the “interrupted” group. Academic performance represents a critical concern for university students. Many students struggling academically may face issues such as absenteeism from classes or failure to submit assignments. When such issues remain unresolved, students may discontinue counseling prematurely. By contrast, this study’s findings highlight the significant impact of sustaining the counselor–student relationship until completion, which not only stabilizes mood by reducing depression and anxiety but also positively influences academic outcomes. Snell [26] demonstrated that students who utilized university-based counseling services experienced symptom relief, increased satisfaction, and improved academic retention. Notably, students who had considered withdrawing from university indicated that counseling played a pivotal role in their decision to remain enrolled. This indicates that counseling services support both mental health and academic persistence, underscoring the responsibility of faculty members in educational and research roles to recognize and understand the efficacy of counseling.

In the short-term subgroup, no statistically significant differences were observed across the eight subscales or within the four critical items. In recent years, Single Session Intervention [27] has gained attention, and studies have shown that even a single session can be effective for addressing depressive and anxiety symptoms in adolescents [28,29]. However, this study suggests that, in Japanese university counseling centers, two sessions are insufficient to produce an effective impact on psychological symptoms. The mid-term group demonstrated significant improvements in depression, generalized anxiety, thought disturbance, and suicidal ideation, while the long-term group showed significant changes in depression, generalized anxiety, and suicidal ideation. The eating concerns subscale had a positive change in the long-term subgroup, suggesting that such issues may not be adequately addressed through counseling alone and may require additional or specialized interventions. Notably, the mean post-treatment scores for depression and generalized anxiety in the mid-term group fell below the clinical cutoff values, whereas such improvements were not observed in the long-term group. These findings suggest that, for Japanese students utilizing university counseling services, mid-term interventions consisting of 3 to 10 sessions may be particularly effective in producing therapeutic change. The relationship between counseling session frequency and therapeutic outcomes has been examined in previous research. Within the framework of the dose–response model, Howard et al. [30] reported that approximately eight sessions are required for 50% of clients to show clinically significant improvement.

Similarly, Hansen et al. [31] estimated that 13 to 18 sessions may be necessary to achieve meaningful therapeutic change. Lambert [32] indicated that while many clients exhibit substantial improvement during the initial sessions, subsequent progress tends to follow a more gradual trajectory. The average number of individual counseling sessions at university counseling centers is approximately 5.9 in the United States [20] and 6.6 in Japan [33].

However, the Center for Collegiate Mental Health emphasizes that the number of sessions should not be uniformly prescribed, but rather flexibly determined based on the severity of clients’ symptoms, therapeutic goals, the resources of the counseling center, students’ preferences and circumstances, and the center’s policies. In this study, data were collected from a Japanese university counseling center that did not impose strict session limits.

Importantly, the findings suggest that students who received a moderate number of sessions (3–10) may have experienced greater therapeutic benefits than those in the short-term (2 sessions) or long-term (11 or more sessions) intervention groups. This represents a noteworthy contribution to the literature, highlighting the value of mid-range counseling interventions. A question warranting future research is the duration of the intervention period. Even with the same total number of sessions (e.g., five), therapeutic timelines differ significantly between weekly and monthly sessions. Thus, considering the temporal spacing of sessions may offer a new and valuable perspective in evaluating counseling effectiveness.

### Limitations

This study has several limitations. First, the data were collected from a single university, which may limit the generalizability of the findings. However, because X University is a moderately sized national general university with relatively balanced student demographics, the findings may still be reasonably generalizable. Future research at institutions with more distinctive demographic characteristics could use these results as a point of comparison.

Second, therapist-related factors, including training, supervision experience, and the type of therapy provided, were not controlled. These variables influence counseling outcomes [34–36]. Nonetheless, all therapists in this study were certified counseling professionals, ensuring a baseline standard of quality. Moreover, counseling sessions were tailored to individual student needs, often combining multiple therapeutic approaches. This flexibility aligns with the study’s objective of assessing the overall effectiveness of therapy in practice rather than evaluating a single standardized intervention.

Third, the number of sessions each student received was not randomized. Session frequency was determined by the students’ needs, reflecting a student-centered counseling approach that allows for self-termination or additional follow-up as appropriate.

Finally, the study lacked a control group, which limits the ability to attribute observed outcomes solely to the intervention. While including a nontreatment or placebo group is the standard method for assessing efficacy, assigning students seeking therapy to such a group would have been ethically inappropriate.

### Future directions

To enhance the robustness of this study, expanding the research design to include multiple universities would improve the generalizability of the findings. Future studies with larger, more diverse samples are warranted to confirm these effects across broader student populations. Additionally, research should examine how different therapy types influence changes in outcome measures. Standardizing administered therapies could facilitate the evaluation of their efficacy, although this may be challenging due to variations in clinical psychologists’ approaches and the dynamic nature of presenting symptoms. Comparing therapy types with session termination or interruption rates could further illuminate students’ preferences for specific therapeutic styles. Collecting data on students who do not complete therapy sessions would provide counseling centers with insights into strategies for improving retention. Finally, follow-up analyses should assess the long-term effects of therapy and whether symptom improvements are sustained over time. Although longitudinal tracking may reduce sample retention, it offers valuable information for both students and university counseling services.

## Conclusion

Therapy is effective for Japanese university students, particularly in alleviating depression, generalized anxiety, thought disturbances, and suicidal ideation. Counseling effects vary according to sex, therapy status, and the number of sessions attended.

Specifically, counseling brings significant improvements in social anxiety among male students and in hostility among female students. Among students who complete therapy, notable reductions are found in academic distress. Interestingly, even students who discontinue therapy show significant improvements in depression and generalized anxiety. Furthermore, a minimum of three sessions are required to achieve measurable therapeutic benefits.

This study contributes to the limited quantitative evidence on counseling outcomes for Japanese university students. It addresses a significant research gap, as few outcome studies have been conducted within university counseling centers in Japan. By doing so, the present study advances academic understanding and supports the accountability of university counseling services by demonstrating the effectiveness and impact of their interventions.

## Data Availability

All relevant data are within the manuscript and its Supporting Information files.

## Acknowledgments

We thank Editage (www.editage.com) for English language editing.

## References

1. Patel V, Prince M. Global mental health: A new global health field comes of age. JAMA. 2010;303: 1976–1977. doi: 10.1001/jama.2010.616.

2. Lipson SK, Zhou S, Abelson S, Heinze J, Jirsa M, Morigney J, et al. Trends in college student mental health and help-seeking by race/ethnicity: Findings from the national healthy minds study, 2013–2021. J Affect Disord. 2022;306: 138–147. doi: 10.1016/j.jad.2022.03.038.

3. Cuijpers P, Smit F, Aalten P, Batelaan N, Klein A, Salemink E, et al. The associations of common psychological problems with mental disorders among college students. Front Psychiatry. 2021;12: Article 573637. doi: 10.3389/fpsyt.2021.573637.

4. Abrams Z. Student mental health is in crisis. Campuses are rethinking their approach. Monit Psychol. 2022;53: 60.

5. Keilson MV, Dworkin FH, Gelso CJ. The effectiveness of time-limited psychotherapy in a university counseling center. J Clin Psychol. 1979;35: 631–636. doi: 10.1002/1097-4679(197907)35:3<631::AID-JCLP2270350329>3.0.CO;2-H.

6. Collins C, Broglia E, Barkham M. Evaluating the evidence base for university counseling services and their effectiveness using CORE measures: A systematic review and meta-analysis. J Affect Disord. 2025;372: 451–462. doi: 10.1016/j.jad.2024.12.022.

7. Chu T, Liu X, Takayanagi S, Matsushita T, Kishimoto H. Association between mental health and academic performance among university undergraduates: The interacting role of lifestyle behaviors. Int J Methods Psychiatr Res. 2023;32: Article e1938. doi: 10.1002/mpr.1938.

8. Duffy A, Keown-Stoneman C, Goodday S, Horrocks J, Lowe M, King N, et al. Predictors of mental health and academic outcomes in first-year university students: Identifying prevention and early-intervention targets. BJPsych Open. 2020;6: Article e46. doi: 10.1192/bjo.2020.24.

9. Horita R, Nishio A, Yamamoto M. Lingering effects of COVID-19 on the mental health of first-year university students in Japan. PLOS One. 2022;17: Article e0262550. doi: 10.1371/journal.pone.0262550.

10. Nagib N, Horita R, Miwa T, Adachi M, Tajirika S, Imamura N, et al. Impact of COVID-19 on the mental health of Japanese university students (years II–IV). Psychiatry Res. 2023;325: Article 115244. doi: 10.1016/j.psychres.2023.115244.

11. An T. A review of international students’ help-seeking. Jpn J Stud Couns. 2022;43: 148–158. doi: 10.57289/jasc.43.2_148.

12. Fujiwara S, Ito R. Changes in major needs of clients of student counseling pre- and post-COVID-19. Jpn J Stud Couns. 2022;43: 171–181. doi: 10.57289/jasc.43.2_171.

13. Egami N, Kawasaki T, Furukawa M, Tanaka T, Keino H, Fujioka I, et al. Counselors’ perceptions about measuring the effectiveness of student counseling services. Jpn J Stud Couns. 2016;37: 94–107.

14. Kasai M. The role of Japanese culture in psychological health: Implications for counseling and clinical psychology. In: Gerstein LH, Heppner PP, Ægisdóttir S, Leung S-MA, Norsworthy KL, editors. International handbook of cross-cultural counseling: Cultural assumptions and practices worldwide. Sage Publications; 2009. pp. 159–171.

15. Packer H. Japan pushes private university mergers as enrolments dwindle; 2025. *Times Book Company Higher Education*. Available from: https://www.timeshighereducation.com/news/japan-pushes-private-university-mergers-enrolments-dwindle.

16. Horita R. Development of the CCAPS Internet-based quick assessment system (CCAPS-iQAS). Jpn J Stud Couns. 2022;43: 182–193. doi: 10.57289/jasc.43.2_182.

17. Horita R, Kawamoto A, Nishio A, Sado T, Locke BD, Yamamoto M. Development of the Counseling Center Assessment of Psychological Symptoms-Japanese version: Pilot study. Clin Psychol Psychother. 2020;27: 97–105. doi: 10.1002/cpp.2412.

18. Horita R, Nishio A, Kawamoto A, Sado T, Locke BD, Yamamoto M. Validity and reliability of the Counseling Center Assessment of Psychological Symptoms-Japanese Version. Jpn Psychol Res. 2023;65: 9–20. doi: 10.1111/jpr.12345.

19. Locke BD, Buzolitz JS, Lei PW, Boswell JF, McAleavey AA, Sevig TD, et al. Development of the counseling center assessment of psychological Symptoms-62 (CCAPS-62). J Couns Psychol. 2011;58: 97–109. doi: 10.1037/a0021282.

20. Center for Collegiate Mental Health. Pennsylvania State University; 2025, 2024 *annual report* (Publication No. STA 25-489). Available from: https://ccmh.psu.edu/assets/docs/CCMH%202024%20Annual%20Report.pdf.

21. McAleavey AA, Nordberg SS, Hayes JA, Castonguay LG, Locke BD, Lockard AJ. Clinical validity of the Counseling Center Assessment of Psychological Symptoms-62 (CCAPS-62): Further evaluation and clinical applications. J Couns Psychol. 2012;59: 575–590. doi: 10.1037/a0029855.

22. Tillman KS, Sell DM, Yates LA, Mueller N. Effectiveness of one-time psychoeducational programming for students with high levels of eating concerns. Eat Behav. 2015;19: 133–138. doi: 10.1016/j.eatbeh.2015.08.002.

23. Wilson GT, Grilo CM, Vitousek KM. Psychological treatment of eating disorders. Am Psychol. 2007;62: 199–216. doi: 10.1037/0003-066X.62.3.199.

24. Lambert MJ. Does client-therapist gender matching influence therapy course or outcome in psychotherapy? Evid Based Med Pract. 2016;2: 1–8. doi: 10.4172/2471-9919.1000108.

25. Mimaki Y, Tsuneyoshi T. On the influence of the relation between gender-role and expectation of acceptance of dependency need on hesitation toward counseling. Bulletin of the Faculty of Education. Educ Psychol, 60, Yamaguchi University. 2010;3, *Arts, Physical Education*: 217–226. Available from: https://cir.nii.ac.jp/crid/1520853834257600640.

26. Snell MN. Treatment effectiveness, client satisfaction, and student retention for university-based counseling services: An outcome and evaluation study. Diss Admin Int A. 1999;60: (6-A), 1925.

27. Hoyt MF. Single session therapy: A clinical introduction to principles and practices. 1st ed. Routledge; 2024. doi: 10.4324/9781003468547.

28. Schleider JL, Weisz JR. Can less be more? The promise (and perils) of single-session youth mental health interventions. Behav Therapist. 2017;40: 256–261. Available from: https://psycnet.apa.org/record/2017-51414-006.

29. Schleider JL, Zapata JP, Rapoport A, Wescott A, Ghosh A, Kaveladze B, et al. Single-session interventions for mental health problems and service engagement: Umbrella review of systematic reviews and meta-analyses. Annu Rev Clin Psychol. 2025;21: 279–303. doi: 10.1146/annurev-clinpsy-081423-025033.

30. Howard KI, Kopta SM, Krause MS, Orlinsky DE. The dose–effect relationship in psychotherapy. Am Psychol. 1986;41: 159–164. doi: 10.1037/0003-066X.41.2.159.

31. Hansen NB, Lambert MJ, Forman EM. The psychotherapy dose–response effect and its implications for treatment delivery services. Clin Psychol: Sci Pract. 2002;9: 329–343. doi: 10.1093/clipsy.9.3.329.

32. Lambert MJ, editor. Bergin and Garfield’s handbook of psychotherapy and behavior change. 6th ed. John Wiley & Sons; 2013.

33. Sugie Y, Sugioka M, Horita R, Fukumori H, Imae H, Kobashi R, et al. Report on the survey about student counseling institutions in 2021. Jpn J Stud Couns. 2022;43: 56–100.

34. Baldwin SA, Wampold BE, Imel ZE. Untangling the alliance–outcome correlation: Exploring the relative importance of therapist and patient variability in the alliance. J Consult Clin Psychol. 2007;75: 842–852. doi: 10.1037/0022-006X.75.6.842.

35. Tracey TJ, Kokotovic AM. Factor structure of the Working Alliance Inventory. Psychol Assess: A J Consult Clin Psychol. 1989;1: 207–210. doi: 10.1037/1040-3590.1.3.207.

36. Wampold BE. How important are the common factors in psychotherapy? An update. World Psychiatry. 2015;14: 270–277. doi: 10.1002/wps.20238.

